# Antigen Specific T Cells in People with Obesity at Five Months Following ChAdOx1 COVID-19 Vaccination

**DOI:** 10.1101/2022.08.16.22278821

**Authors:** Neil E. Wrigley Kelly, Conor De Barra, Ferrah Shaamile, Aisling Holland, Liam Shaw, Patrick W.G. Mallon, Jean O’Connell, Andrew E. Hogan, Donal O’Shea

## Abstract

**Background/Objectives:** People with obesity (PWO) face an increased risk of severe outcomes from COVID-19, including hospitalisation, ICU admission and death. Obesity has been seen to impair immune memory following vaccination against influenza, hepatitis B, tetanus, and rabies. Little is known regarding immune memory in PWO following COVID-19 adenovirus vector vaccination.

**Subjects/Methods:** We investigated SARS-CoV-2 specific T cell responses in 50 subjects, five months following a two-dose primary course of ChAdOx1 nCoV-19 (AZD1222) vaccination. We further divided our cohort into PWO (n=30) and matched controls (n=20). T cell (CD4^+^, CD8^+^) cytokine responses (IFNγ, TNFα) to SARS-CoV-2 spike peptide pools were determined using multicolour flow cytometry.

**Results:** Circulating T cells specific for SARS-CoV-2 were readily detected across our cohort, with robust responses to spike peptide stimulation across both T cell lines. PWO and controls had comparable levels of both CD4^+^ and CD8^+^ SARS-CoV-2 spike specific T cells. Polyfunctional T cells – associated with enhanced protection against viral infection – were detected at similar frequencies in both PWO and controls.

**Conclusions:** These data indicate that PWO who have completed a primary course of ChAdOx1 COVID-19 vaccination have robust, durable, and functional antigen specific T cell immunity that is comparable to that seen in people without obesity.

## Introduction

People with obesity (PWO) have a substantially increased risk of developing severe coronavirus disease 2019 (COVID-19). Many studies have summarised this relationship (1, 2). One meta-analysis for example outlined PWO to have a 113% increased risk of hospitalisation, 74% increased ICU admission and 48% higher mortality (1). The introduction of multiple highly efficacious COVID-19 vaccines mitigates the impact of the pandemic for many. This is of particular importance to those with obesity. However, little is known regarding immune memory following COVID-19 vaccination in this population.

PWO have impaired responses to vaccination in multiple settings. Impaired humoral responses have been observed following rabies (3), tetanus (4) and hepatitis B (5) immunisation. Furthermore, a study published in 2012 looked at PWO and controls without obesity following influenza vaccination, showing PWO to have both greater declines in antibody titres and lower frequencies of antigen specific CD8^+^ T cells at 12 months follow-up (6).

Antigen specific T cells have a crucial role in the adaptive response to SARS-CoV-2 infection and are protective against severe disease (7). They are durable, with longitudinal cohorts demonstrating their presence for at least 8 months post-vaccination (8). They are also more resilient to antigenic variation than antibody/B cell responses, as observed in the case of the omicron variant of concern (9).

The aim of this study was to evaluate if adults (18 years and older) with obesity (body mass index, BMI ≥ 30.0 kg/m^2^) who have a history of COVID-19 adenovirus vector vaccination demonstrate poorer antigen specific T cell immunity compared to controls.

## Materials/Subjects and Methods

We performed a cross-sectional cohort study looking at SARS-CoV-2 specific CD4^+^ and CD8^+^ cell responses at 5 months following completion of a two-dose course of ChAdOx1 nCoV-19 (AZD1222) vaccine, with a dosing interval of 12 weeks. Subjects were recruited from staff and outpatients attending the St. Columcille’s Hospital, Loughlinstown COVID-19 vaccination centre. 50 volunteers (30 PWO [BMI ≥30 kg/m^2^] and 20 controls [BMI <30 kg/m^2^]) were included (see Table 1). Samples were collected over November and December 2021. Inclusion criteria were: ability to give informed consent, age ≥18 years, 4 to 6 months since second dose of ChAdOx1 nCoV-19 (AZD1222) vaccine. Exclusion criteria included: known previous SARS-CoV-2 infection (confirmed by PCR or antigen test), treatment with certain classes of immunosuppressants (anti-TNF agents, IL-1, IL-2, IL-6, IL-17 inhibitors) within the preceding 8 months, obesity induced by other endocrine disorders, active lymphoproliferative disorder and any active illness or medication which in the opinion of the investigator precluded participation in the study.

**Table 1:** Participant Characteristics

**Figure 1**
| Table 1: Participant Characteristics |  |  |
| --- | --- | --- |
|  | People with Obesity | Controls |
| Total number (%) | 30 (60) | 20 (40) |
| Age (years) | 60.5 (47.5-67.0) | 63.0 (60.3-65.0) |
| BMI (kg/m<sup>2</sup>) | 39.1 (32.6-47.0) | 26 (22.8-27.8) |
| Female (n [%]) | 18 (60) | 8 (40) |
| Days from second vaccine dose | 158 (153-166) | 154 (152-158) |
| *Data are median (IQR) unless specified. BMI: body mass index.* |  |  |

Subjects were recruited as part of the Obesity Immunology Group cohort study. All subjects provided written, informed consent. The study was conducted in accordance with the Declaration of Helsinki and ethical approval was granted by the St. Vincent’s Hospital Group Research Ethics Committee.

### Preparation of peripheral blood mononuclear cells (PBMC) and flow cytometric analysis

PBMCs were isolated from whole blood samples by centrifugation through density gradient media (Lymphoprep™, Nycomed Pharma, Oslo, Norway), before suspension in cell cryopreservation media and storage in a -70 °C freezer.

Thawed PBMCs were stimulated with SARS-CoV-2 spike peptide pools (Miltenyi PepTivator^®^SARS-CoV-2 research grade). After 4 hours protein transport inhibitor (Thermo Fisher Scientific, Waltham, Massachusetts, USA) was added to each well, samples were incubated at 37 °C for a further 12 hours. Having completed incubation, cells were centrifuged, re-suspended in flow cytometry staining buffer, then stained for cell surface markers (CD3, CD4, CD8). Following staining, live cells were fixed, permeabilised and intracellular cytokine staining was performed. Multicolour flow cytometry was performed using Attune™ NxT Flow Cytometer.

### Quantification and statistical analysis

Data and statistical analyses were performed using FlowJo v10.8.1 and GraphPad Prism 9 software. Statistical comparisons were performed using the Wilcoxon and Mann-Whitney U test, where appropriate. Statistical significance for p values was <0.05. Background subtraction was used when calculating antigen specific T cell frequencies.

## Results

The clinical characteristics of the cohorts are presented in table 1.

### Robust SARS-CoV-2 specific T cell responses in people with obesity

Adenovirus vector vaccines have been shown to elicit robust, durable SARS-CoV-2 specific T cell responses following completion of vaccine course (8). We confirmed this to be the case in our cohort, where both CD4^+^ and CD8^+^ cells were seen to express interferon gamma (IFNγ) following stimulation with spike peptide pools (Figure 1A-C) (median CD8^+^ IFNγ^+^ frequency following spike stimulation: 0.780%, IQR 0.451-1.235%; median CD4^+^: 0.028%, IQR 0.013-0.051%).

**Figure 1.**
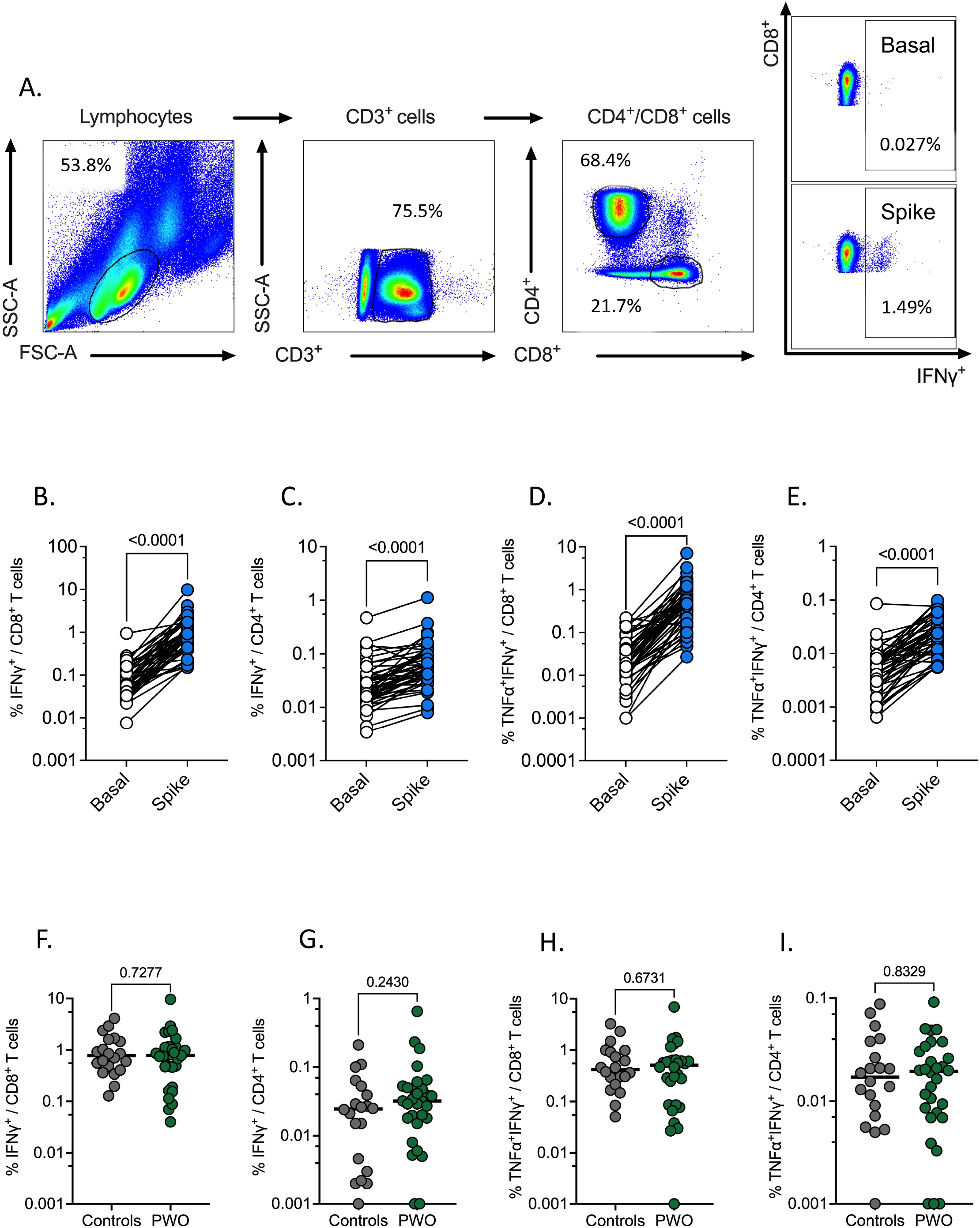
SARS-CoV-2 spike specific T cell responses of subjects at 5 months following ChAdOx1 COVID-19 vaccination. (A) Representative flow cytometry gating strategy for detection of T cell subsets (CD8^+^ or CD4^+^) and their cytokine production. (B-E) Scatter plot showing the frequencies of cytokine expressing T cells (CD8^+^ or CD4^+^) from vaccinated individuals following overnight stimulation with SARS-CoV-2 spike peptide pools. (F-I) Frequencies of T cell subsets (CD8^+^ or CD4^+^) in COVID-19 vaccinated individuals with (PWO) or without obesity (controls), producing cytokine (TNFα, IFNγ) in response to overnight stimulation with SARS-CoV-2 spike peptide pools. Where bars are present they denote median. Statistical comparisons across cohorts were performed with the Wilcoxon test for paired data (B-E) and Mann-Whitney U test for unpaired data (F-I). Data for (F-I) were background subtracted against negative control.

T cells that produce multiple cytokines are known as polyfunctional T cells. Their presence among the antigen specific T cell populations elicited by vaccination is highly desirable, as they are associated with enhanced protection against viral infections (10). Among our cohort polyfunctional T cells (IFNγ^+^TNFα^+^) were readily detectable across both conventional (CD4^+^ and CD8^+^) T cell lines following spike stimulation (Figure 1D-E) (median CD8^+^ IFNγ^+^TNFα^+^ frequency against spike: 0.463%, IQR 0.203-0.783%; median CD4^+^: 0.014%, IQR 0.008-0.031%).

Obesity has been associated with substantial innate and adaptive immune dysregulation, including impaired anti-viral immunity following infection or vaccination (6, 11). To investigate this, we separated our cohort into PWO and controls (median BMI 39.1 kg/m^2^ [IQR 32.6-47.0] and 26 kg/m^2^ [IQR 22.8-27.8]). PWO were seen to have robust SARS-CoV-2 specific T cell responses, with no differences in the frequencies of CD4^+^ and CD8^+^ T cells producing IFNγ in response to spike stimulation (Figure 1F-G). Furthermore, similar frequencies of polyfunctional T cells were observed in those with and without obesity (Figure 1H-I).

## Discussion

Given the greatly increased risk of severe COVID-19 in people with obesity it is important to properly characterise vaccine induced immunity in this vulnerable population. Here we studied antigen specific T cell responses in PWO and controls persisting to 5 months following a complete primary course of ChAdOx1 nCoV-19 (AZD1222) vaccine. Robust frequencies of antigen specific CD4^+^ and CD8^+^ T cells were identified across our cohort (n=50). Between group analysis provides evidence that SARS-CoV-2 specific T cell frequencies are preserved in PWO.

Specific forms of immunosuppression have been associated with impaired responses to COVID-19 vaccines. These include people with haematological malignancies (12), solid organ transplant recipients (13) and people with advanced HIV infection (14). Obesity has been associated with extensive immune dysregulation, with chronic systemic inflammation and loss of host protection (15). PWO have been seen to have impaired immune memory in multiple settings. Weakened antibody responses have been seen following rabies (3), tetanus (4) and hepatitis B vaccination (5). While impairment of both humoral and CD8^+^ T cell responses has been seen following influenza vaccination (6).

Many questions remain regarding immune memory in PWO following COVID-19 vaccination. Most of the available evidence relates to the mRNA vaccines. The BNT162b2 vaccine has been associated with impairment of antibody responses in abdominal obesity (16). While follow-up of the phase III mRNA-1273 and BNT162b2 studies - at 5 and 6 months respectively - showed very similar vaccine efficacy against symptomatic infection between people with and without obesity (17, 18). Regarding COVID-19 adenovirus vector vaccines, a study by Lee et al. showed antibody (IgG against receptor binding domain) responses to the ChAdOx1 nCoV-19 vaccine to be unimpacted by elevated BMI (19). However, there are no published papers describing antigen specific T cell responses in PWO following COVID-19 adenovirus vector vaccination.

Our data demonstrate that PWO who have completed a two-dose course of ChAdOx1 nCoV-19 vaccine have robust antigen specific T cell responses along both CD8^+^ and CD4^+^ lines. Responses that are comparable in frequency to controls, as well as being comparable to the COVID-19 vaccine literature in general (8). The abundant presence of IFNγ expressing cells implies not just antigen recognition but functional capacity to inhibit viral replication and mobilise the immune response to infection. Polyfunctional T cells – readily detectable in PWO - have been seen to correlate with favourable disease evolution and protection (10).

As SARS-CoV-2 reaches a point of endemicity with primary vaccine courses, boosters, infections, and not infrequent reinfections, it is important for us to understand the implications of each antigenic exposure for vulnerable hosts. Physiological contraction of peripheral antibody titres following infection (20) and vaccination (8), as well as viral antigenic variation leading to immune escape diminish the protection conferred by humoral immunity. This reinforces the importance of cellular (antigen specific T cell, memory B cell) immunity, particularly in view of the evidence for conservation of T responses to the antigenic variation thus far observed in variants of concern (9).

Our study has limitations. While the groups in this study were well matched for age and mean days from second vaccine dose, they were less well matched for sex. Future research regarding immune memory in PWO in COVID-19 is critical. As the pandemic progresses, longitudinal studies are required in order to continue to characterise the longevity of cellular and humoral responses in this population, as well as to define precise correlates of protection.

In summary, we measured SARS-CoV-2 specific CD4^+^ and CD8^+^ cell responses in people with obesity following ChAdOx1 COVID-19 vaccination, showing features associated with protective immunity and providing evidence that antigen specific T cell frequencies are not impacted by obesity.

## Data Availability

All data produced in the present study are available upon reasonable request to the authors

## Acknowledgements

The authors wish to thank all study participants and their families. We would like to thank Prof. Alan Landay and Prof. Michael Turner, for their expert input and support. We wish to acknowledge the Irish Coalition for People Living with Obesity (ICPO) for their important advocacy work.

## Author Contributions

NEWK optimized the experimental protocol and performed the experiments. CDB provided technical assistance. NEWK, FS, AH, LS, JOC and DOS recruited study participants. NEWK processed and biobanked samples. NEWK, JOC and DOS analysed the clinical cohorts. NEWK and AEH analysed the data and drafted the manuscript. AEH, DOS, JOC, PM and NEWK conceptualized/designed the study and approved the final manuscript as submitted.

## Data availability statement

The datasets generated during and/or analysed during the current study are available from the corresponding author on reasonable request.

